# Active methods, and their educational strategies during residencies of health professions: a scope review

**DOI:** 10.1101/2023.02.21.23286249

**Authors:** Vera Lúcia Garcia, Everton Soeiro, José Maurício de Oliveira, Laura Schiesari, Romeu Gomes, Rachel Riera

**Author notes:** How to cite: Garcia VL, Soeiro E, Oliveira JM, Schiesari L, Gomes, R & Riera R. (2023, [02] [21]). Active methods, and their educational strategies during health residencies: a scope review. MEdRxiv (medRxiv Manuscript Processing System).

## Abstract

The objective of this study is to map and evaluate the scientific production, on the different strategies / active methods in the context of residences of health professions. The study will be developed at Hospital Sírio-Libanês, São Paulo, Brazil. The scope review will be developed based on the methodological framework of the Joanna Brigss Institute (Peters et al., 2017). The review report will follow recommendations of the Preferred Reporting Items for Systematic Reviews and Meta Analyses extension for scoping reviews (PRISMA ScR) (Tricco, 2018). The protocol for this review was planned and made available on the MEdRxiv preprint database prior to the start of conducting the review. The following research question will guide for searching in databases: What and how are the different active methods used in the context of residencies of health professions? A broad and sensitive search will be carried out in the literature through structured search strategies, with relevant descriptors and synonyms, for the following databases or data repositories in the areas of education and/or health listed below: Biblioteca Virtual de Saúde (BVS), British Education Index (BEI), Campbell Collaboration, Cochrane Library (via Wiley), Education Research Complete (via EBSCO), Educational Resources Information Center – ERIC, Excerpta Medica Database (EMBASE, via Elsevier), Medical Literature Analysis and Retrieval System Online (MEDLINE, via PubMed), Online Education Database.

## 1. Introduction

New challenges are required in the current scenarios of practice, especially in residency for health professions, which care, and training are integrated. To meet social demands, transformations in the education of health professionals and new ways of working with knowledge were required. This demand gives way to the growing tendency to seek innovative methods, which admit an ethical, critical, reflexive, and transformative pedagogical practice, going beyond the limits of purely technical training, to effectively achieve training (Mitre et al., 2008).

John Dewey and Jerome Bruner are precursors of problem-based learning (PBL). The thought of the American philosopher Dewey was marked by ideas of philosophical pragmatism and believed that philosophy should approach the universe of daily life in a practical, pragmatic way, replacing dogmatism with the experimental method, and until then, knowledge was seen in isolation, without useful meaning (Dewey, 2011). The first problem-based curriculum organization used in the medical course curriculum at McMaster University, Canada, in the late 1960s - employed ideas from Dewey and Bruner.

According to Lima (2017), in recent decades, in addition to PBL, other methodologies have been discussed, such as “problematization, and project-based learning, in teams, through games or the use of simulations” (p. 424). Also, according to the author, the so-called active methods “are considered technologies that provide engagement of students in the educational process and that favor the development of their critical and reflective capacity in relation to what they are doing” (Lima, 2017, p. 424).

An educational strategy refers to the art of applying the available means to achieve specific educational objectives (Anatasiou, Alves, 2012). In this way, the educator must plan and use the best tools so that students can develop their skills. Educational strategies make up a framework of tools that can be used in different curricula. The active methods go through the curriculum, and it is explained in the pedagogical project of the course/residency program.

## 2. Justification

Considering the broad discussion that has been developed in the field of health education about active methods, it is necessary to map, through a scope review, to identify the nature and extent of the literature related to the key concepts of these educational strategies/methodologies, to identify the educational strategies/methodologies used in the context residences of health professions and gaps in the literature.

## 3. Objective

Map and evaluate the scientific production, on the different strategies / active methods in the context of residences of health professions.

## 4. Methods

The study will be developed at Hospital Sírio-Libanês, São Paulo, Brazil. The scope review will be developed based on the methodological framework of the Joanna Brigss Institute (Peters et al., 2017). The review report will follow recommendations of the Preferred Reporting Items for Systematic Reviews and Meta Analyses extension for scoping reviews (PRISMA ScR) (Tricco, 2018). The protocol for this review was planned and made available on the MEdRxiv preprint database prior to the start of conducting the review.

### 4.1 Research question(s)

Stakeholders were consulted throughout the development of this review protocol aiming of increasing the applicability of its results and supporting their communication and translation enabling their use by society. The following stakeholders were consulted: consumers (managers, professor/preceptors, and students), specialists in active methods, specialists in scope review methodology.

The research question for this review was structured using the acronym PCC as follows:

- P (population, condition): health educators, managers, or residents of health professions.
- C (concept): active methods, innovative methodologies.
- C (context): mandatory, elective, or optional / practical or theoretical / face-to-face, remote or hybrid teaching activities. (Traditional, PBL etc.).

The following research questions will guide for searching in databases: What and how are the different active methods used in the context of residencies of health professions?

### 4.2 Inclusion and exclusion criteria

- P (population, condition): health educators, managers, or residents of health professions.
- C (concept): active methods, innovative methodologies. Scientific evidence is obtained from the results of scientific studies and used to support or refute a recommendation. Thus, strategies were considered with the objective of translating scientific and/ or methodological information into format/content aimed at ensuring the understanding of terms, criteria, tools, and approaches related to scientific evidence.
- C (context): any strategy related to the communication of scientific evidence in health was considered, at the individual or population level, within the scope of public or supplementary health, at any level of assistance (health unit, neighborhood, municipality, state, region, or country).

Primary and secondary studies will be considered. No language filter will be applied, addressing issues related active methods of residencies of health professions. Studies that are related to contexts other than residencies will be excluded.

The search will be restricted to the period from the year 2000 considering the advances and changes related to residencies.

### 4.3 Search strategies

A broad and sensitive search will be carried out in the literature through structured search strategies, with relevant descriptors and synonyms, for the following databases or data repositories in the areas of education and/or health listed below: Biblioteca Virtual de Saúde (BVS), British Education Index (BEI), Campbell Collaboration, Cochrane Library (via Wiley), Education Research Complete (via EBSCO), Educational Resources Information Center – ERIC, Excerpta Medica data BASE (EMBASE, via Elsevier), Medical Literature Analysis and Retrieval System Online (MEDLINE, via PubMed), Online Education Database (https://www.oecd.org/education/database.htm).

Additional unstructured searches will be performed on the following sources related to education or health systems: American Educational Research Association (AERA); Association for Medical Education in Europe (AMEE); Best Evidence Medical Education (BEME); Cochrane Effective Practice and Organization of Care (EPOC) (https://epoc.cochrane.org/); European Association for Research on Learning and Instruction (EARLI), Institute of Education Sciences (https://eric.ed.gov/); Joanna Briggs Institute (https://jbi.global/).

Other additional searches will be conducted by consulting the reference lists of relevant studies and contacting experts in the field. Full publications, abstracts presented at congresses and events, online reports, theses, and dissertations will be included.

### 4.4 Selection of evidence sources

After the exclusion of duplicates, the selection of references will be conducted in a two-step process using Rayyan platform [Ouzzani et al., 2016]. At the first phase, a pair of reviewers will independently assess titles and abstracts retrieved. References classified as ‘potentially eligible’ will be screened at the second phase, by the reading of the full text to confirm eligibility. Divergences between reviewers will be solved by consulting a third reviewer. References excluded after the second phase will be presented in the ‘excluded studies table’ along with the reasons for each exclusion.

### 4.5 Data extraction

Three reviewers will conduct calibration of data extraction, independently, contemplating how many articles are necessary to reach homogeneity of the process. Subsequently, two reviewers will extract data, in a complementary way. A spreadsheet for extraction will be created in Microsoft Excel®, containing the following information: author, year of publication, institution, financial support, study design, location, number of participants, participant characteristics, issues or outcomes analyzed, results, limitations, knowledge gaps. The following data were collected for each identified strategy:

1. Main category: active methods, innovative methodology
2. Target audience: residents of health professions – medical or non-medical.
3. Name of the innovative strategy or methodology.
4. Duration of the strategy: continuous or sporadic.
5. *Status* of the strategy: proposed, implemented, and not evaluated, implemented, and evaluated.
6. Impacts expected by the proponents (outcomes).
7. Barriers and facilitators identified for implementation.

### 4.6 Analysis of evidence and presentation of results

The extracted data will be analyzed first in a descriptive way, to characterize scientific production on the subject. Next, the data will be explored through an adaptation of the content analysis technique (Gomes, 2007), thematic modality, described by Bardin (1979). Results will be presented in a descriptive way and through tables, figures and graphs that express the results properly.

Assessment of the methodological quality of the included studies will not be conducted, being considered optional in the scoping review (Peters et al, 2017). This qualitative synthesis will be presented using a narrative approach and in graphs and/or tables. Depending on the availability of information, descriptive statistics will be performed using Microsoft Excel® and/or STATA®.

## Data Availability

The protocol for this review was planned and made available on the MEdRxiv preprint database prior to the start of conducting the review.

